# Post-COVID conditions during Delta and early-Omicron SARS-CoV-2 variant periods among adults in the United States

**DOI:** 10.1101/2023.08.09.23293776

**Authors:** Deja Edwards, Pamela Logan, Leora R. Feldstein, Tarayn Fairlie, Emma Accorsi, Sharon Saydah

## Abstract

**Background:** Post-COVID conditions after infection with new SARS-CoV-2 variants have been incompletely described. We compared the prevalence and risk factors for ongoing symptoms lasting 4 weeks or longer (often referred to as post-COVID Conditions) among adults who had tested positive vs. negative during the Delta and early-Omicron periods.

**Methods:** Self-reported survey data regarding symptoms and previous SARS-CoV 2 test results were collected from May 31 – July 6, 2022, from a probability sampling of United States adults. Respondents were classified according to their test result, predominant circulating variant when respondents first tested positive (Delta vs early-Omicron), and demographic risk factors.

**Results:** Among 2,421 respondents, 256 tested positive during Delta, 460 during early-Omicron, and 1,705 always tested negative. Nearly one-fourth (22.3%) of negative respondents reported ≥1symptom that lasted ≥4 weeks, compared to 60.6% (p<0.05) of respondents who tested positive during the Delta period and 47.8% (p<0.05) during the early-Omicron period. Fatigue, change in smell/taste, and cough were commonly reported by respondents who tested positive. Demographic risk factors associated with ongoing symptoms were being female and unemployed (aOR 1.28, 95% CI 1.06–1.55; aOR 1.48, 95% CI: 1.17–1.87).

**Conclusion:** The reported occurrence of ongoing symptoms associated with post-COVID conditions was reduced during the early-Omicron period, compared with Delta.

## Introduction

Ongoing symptoms have been reported by a growing number of adults previously infected with SARS-CoV-2. These ongoing symptoms, lasting four or more weeks, have been referred to as *post-COVID conditions* or *long COVID*^1^. The U.S. Census Bureau’s Household Pulse Survey estimates 14.7% of adults have experienced an ongoing symptom three or more months after infection^2^. Previous studies suggest post-COVID conditions are more likely to occur among people with more severe acute COVID-19 illness, persons with underlying conditions, and those who did not receive the COVID-19 vaccine^1,3^. Older adults and females have also reported a higher occurrence of post-COVID conditions^4,5^. Common post-COVID conditions reported by patients include fatigue, shortness of breath, and cognitive dysfunction^6,7,4,8^. Many studies researching post-COVID conditions use data from electronic health records. Using survey data allows us to capture patient reported symptoms, including those among patients who were non-hospitalized, asymptomatic, or had less severe symptoms.

Few researchers have investigated whether post-COVID conditions differ by variant period. Some evidence suggests the occurrence of post-COVID conditions is lower among those infected during the Omicron period compared to Delta. Reasons for these observed differences in occurrence could include milder cases of acute COVID-19 among those infected with the Omicron variant or higher rates of vaccination prior to infection among those infected with the Omicron variant^9,10,11,12, 13,14,15^.

In the current study, we estimated the prevalence of ongoing symptoms associated with first testing positive for SARS-CoV-2 during the Delta and early-Omicron variant periods and among adults testing negative using a nationwide survey of adults in the U.S.

## Methods

### Study Design & Study Sample

We conducted analyses using cross-sectional data collected by Porter Novelli (PN) Public Services^16^ using PN SummerStyles 2022, a nation-wide survey of U.S. adults administered from May 31, 2022 – July 6, 2022. PN SummerStyles 2022 respondents were selected from a sample of approximately 6,000 panel members who were 18 years and older. The survey was administered by the market research firm Ipsos via their KnowledgePanel, a continuously replenished panel of approximately 60,000 panelists who are representative of the non-institutionalized U.S. population. Panelists were randomly recruited by mail using probability-based sampling by address, regardless of whether the household had a landline telephone or internet access. A laptop and internet access were provided, if needed, for survey panelists. Participation was voluntary, and respondents were able to skip questions or discontinue participation with the survey or panel at any time. Respondents received cash-equivalent reward points as compensation for participating.

We analyzed responses of deidentified respondents who reported being tested for SARS-CoV-2 infection. For this analysis, we used statistical weighting to align the sample with U.S. population distributions, adjusting for gender, age, and education. These recommended weights were provided by Porter Novelli Public Services which were designed to match the U.S. Current Population Survey (CPS) proportions^16^. For data currency, we used the most recent weighting from U.S. Census’ American Community Survey (ACS) data^17^.

### Variable Definitions

#### Socio-demographics

Respondents reported their age, sex, race/ethnicity, income, household size, highest level of education, marital status, community type, census region, and employment status. Poverty level was determined by comparing the reported household income to the poverty threshold value provided by the U.S. Census Bureau^17^, based on respondent’s household size.

#### SARS-CoV-2 Test Status

Respondents reported ever having received a positive SARS-CoV-2 test result (“I tested positive at least once for COVID-19”), always receiving a negative SARS-CoV-2 test result (“I have been tested and my results have always been negative”) or never having been tested for the infection (“I have never been tested”). Those who reported having received a positive or negative result were included in the analysis. Respondents were then grouped based on their SARS-CoV-2 test status. Respondents who reported never receiving a positive test comprised the *negative* group. Those who reported a positive test were separated into groups by SARS-CoV-2 variant period based on the self-reported date of their first positive test. Variant predominant periods were defined based on the CDC’s COVID Data Tracker^18^—pre-Delta variants included those reporting testing positive before July 2021, Delta from July 2021 through December 2021 (Delta positive), and early-Omicron from January 2022 to June 2022 (early-Omicron positive), when the survey was completed. Only respondents who reported testing negative, first testing positive during the period when the Delta variant was predominant, or first testing positive when early-Omicron was the predominant variant were included in the analysis.

#### Symptoms

Respondents who reported testing positive for SARS-CoV-2 infection were asked whether, following their positive test, they experienced 1 or more of 17 symptoms that lasted for four weeks or longer (See Appendix—Symptoms List). Those who tested negative were asked whether, in the past month, they experienced any of the same symptoms lasting longer than four weeks. Symptoms were also grouped into bodily systems they most often affect based on information from previous studies^4^ – gastrointestinal, neurological, respiratory/cardiac, and other.

#### COVID-19 Vaccination Status

Respondents were asked whether they had completed a COVID-19 vaccine primary series (received two doses of a two-dose series or one dose of a single dose series, information on additional doses, and/or booster doses was not collected). Respondents were also asked the date of their most recent vaccination. We used both dates to determine whether respondents were vaccinated (either tested positive ≥30 days after completing vaccination series or completed primary series ≥30 days prior to completing survey for those testing negative).

#### Healthcare Utilization

Respondents were asked whether they sought healthcare services for their symptoms that lasted longer than 4 weeks since they first experienced the symptom(s). Responses were categorized as receiving outpatient care (“reported seeing a doctor, nurse, or other health professional or going to urgent or emergency care”); receiving hospital-based care (“reported being hospitalized, treated in the intensive care unit, or being given a breathing tube and ventilator); or receiving no healthcare services (“reported none of these”).

#### Analytic Sample

Of the 5,990 individuals invited to participate in the SummerStyles survey, 4,156 adults completed the survey (response rate 69.4%). We excluded adults who: 1) did not answer whether they were ever tested for SARS-CoV-2 (n=23), 2) indicated they were never tested for SARS-CoV-2 (n=1210), 3) skipped questions about ongoing symptoms (n=61), or 4) tested positive during a pre-Delta period (n=441). Data from 2,421 adults were included in analysis.

### Statistical Analysis

To determine if respondent characteristics and report of ongoing symptoms differed by SARS-CoV-2 variant and test status, we compared the following groups: Delta positive to early-Omicron positive, Delta positive to negative, and early-Omicron positive to negative. To determine if respondent characteristics and reports of ongoing symptoms differed by SARS-CoV-2 variant and test status, we used Wald Chi-square tests to compare the following groups: testing positive during Delta to testing positive during early-Omicron, testing positive during Delta to negative, and testing positive during early-Omicron to negative. Using multivariable logistic regression models, we estimated the odds ratio with 95% confidence intervals (CI) of having at least one ongoing symptom lasting four weeks or longer by test status. The first model assessed the odds of reporting ongoing symptoms among all respondents. We then restricted the logistic regression model to only adults reporting a positive test to determine potential factors associated with ongoing symptoms among respondents who reported testing positive. These models were adjusted for variant predominance period, age (18-39, 40-59, and 60 years and older), sex, and employment status (working full-time, working part-time, not working) as potential confounders. Statistical significance was defined as a p-value < 0.05.

All analyses were conducted using RStudio, version 2022.07.1. Statistical weighting was used to align the sample with the noninstitutionalized U.S. population distributions, accounting for gender, age, household income, race/ethnicity, household size, education, census region, and metropolitan status (i.e., urban/rural differences) using weights provided by Porter Novelli (PN)^16^.

### Human subjects protection

This activity was reviewed by CDC and was conducted consistent with applicable federal law and CDC policy. (See e.g., 45 C.F R. part 46, 21 C.F R. part 56; 42 U S C. x241(d); 5 U S C. x551a; 44 U S C. x3501 et seq.) CDC licensed this data from Porter Novelli Public Services. While Porter Novelli Public Services and its vendors are not subject to CDC Human Subject Review, they adhere to all professional standards and codes of conduct set forth by the Council of American Survey Research Organizations (CASRO). Respondents were informed that their answers are being used for market research and they may refuse to answer any question at any time. No personal identifiers are included in the data file that is provided to CDC.

## Results

### Respondent Characteristics

More than two-thirds of respondents reported always testing negative for SARS-CoV-2 (n=1705). Among those who tested positive at least once, the majority first tested positive during the early-Omicron period (n=460), and fewer first tested positive during the Delta period (n=256) (Table 1).

**Table 1.** Characteristics of study respondents by testing status, weighted°.

|  | First testing<br>positive during<br>Delta | 95% CI | First testing<br>positive during<br>Omicron | 95% CI | Report<br>only<br>testing<br>negative | 95% CI | Delta v.<br>Omicron | Delta v.<br>Negative | Omicron v.<br>Negative |
| --- | --- | --- | --- | --- | --- | --- | --- | --- | --- |
|  | n = 257 |  | n = 460 |  | n = 1,705 |  |  |  |  |
| **Age group** |  |  |  |  |  |  | **0.4** | **<0.001** | **<0.001** |
| 18-39 | 44.1% | 37.8% - 50.7% | 38.3% | 33.1% - 43.4% | 32.3% | 29.6% - 35.0% |  |  |  |
| 40-59 | 34.5% | 28.1% - 41.0% | 36.5% | 31.5% - 41.8% | 29.9% | 27.2% - 32.6% |  |  |  |
| >= 60 | 21.4% | 15.0% - 27.9% | 25.3% | 20.1% - 30.4% | 37.9% | 35.2% - 40.6% |  |  |  |
| **Sex** |  |  |  |  |  |  | **0.7** | **0.3** | **0.07** |
| Male | 45.5% | 39.6% - 51.8% | 43.7% | 39.1% - 48.8% | 49.5% | 47.0% - 52.0% |  |  |  |
| Female | 54.5% | 48.5% - 60.8% | 56.3% | 51.6% - 61.3% | 50.5% | 48.0% - 53.1% |  |  |  |
| **Marital status** |  |  |  |  |  |  | **0.5** | **0.4** | **>0.9** |
| Married | 53.6% | 47.6% - 60.0% | 59.2% | 54.6% - 64.1% | 59.1% | 56.6% - 61.6% |  |  |  |
| Single | 31.6% | 25.7% - 38.1% | 26.8% | 22.2% - 31.7% | 26.5% | 24.1% - 29.0% |  |  |  |
| Other | 14.8% | 8.8% - 21.3% | 14.0% | 9.3% - 18.9% | 14.4% | 12.0% - 16.9% |  |  |  |
| **Highest level of**<br>**education** |  |  |  |  |  |  | **0.012** | **0.13** | **0.006** |
| Less than HS | 11.7% | 5.3% - 18.2% | 2.7% | 0% - 7.7% | 7.7% | 5.1% - 10.3% |  |  |  |
| HS | 29.8% | 23.5% - 36.4% | 24.2% | 19.2% - 29.4% | 24.8% | 22.3% - 27.5% |  |  |  |
| Some college | 26.1% | 19.7% - 32.6% | 29.8% | 24.7% - 34.9% | 26.4% | 23.8% - 29.1% |  |  |  |
| Bachelors or higher | 32.5% | 26.1% - 39.0% | 43.3% | 38.2% - 48.3% | 41.1% | 38.5% - 43.8% |  |  |  |

| **Race/Ethnicity** |  |  |  |  |  |  | **0.3** | **0.003** | **0.2** |
| --- | --- | --- | --- | --- | --- | --- | --- | --- | --- |
| White, non-Hispanic | 68.2% | 62.9% - 73.9% | 63.2% | 58.8% - 67.8% | 62.6% | 60.2% - 64.9% |  |  |  |
| Black, non-Hispanic | 7.0% | 1.8% - 12.8% | 9.7% | 5.3% - 14.3% | 12.8% | 10.5% - 15.2% |  |  |  |
| Other, non-Hispanic | 3.1% | 0% - 8.9% | 6.9% | 2.5% - 11.5% | 8.9% | 6.5% - 11.2% |  |  |  |
| Hispanic | 20.4% | 15.2% - 26.2% | 18.2% | 13.8% - 22.8% | 14.4% | 12.1% - 16.7% |  |  |  |
| 2+ races, non-Hispanic | 1.3% | 0% - 7.1% | 2.1% | 0% - 6.7% | 1.4% | 0% - 3.7% |  |  |  |
| **Community type** |  |  |  |  |  |  | **0.5** | **0.5** | **0.7** |
| Urban | 39.4% | 32.9% - 45.7% | 36.3% | 31.5% - 41.4% | 35.0% | 32.5% - 37.7% |  |  |  |
| Rural | 16.2% | 9.8% - 22.6% | 14.0% | 9.1% - 19.1% | 15.7% | 13.1% - 18.3% |  |  |  |
| Suburban | 44.4% | 38.2% - 50.9% | 49.7% | 44.9% - 54.8% | 49.3% | 46.7% - 51.9% |  |  |  |
| **Census region** |  |  |  |  |  |  | **0.015** | **0.006** | **>0.9** |
| Northeast | 16.6% | 10.6% - 23.3% | 19.2% | 14.4% - 24.4% | 18.4% | 15.8% - 21.0% |  |  |  |
| Midwest | 29.5% | 23.5% - 36.2% | 17.2% | 12.4% - 22.4% | 18.1% | 15.5% - 20.7% |  |  |  |
| South | 36.7% | 30.7% - 43.4% | 38.1% | 33.1% - 43.2% | 37.0% | 34.4% - 39.6% |  |  |  |
| West | 17.3% | 11.3% - 24.0% | 25.5% | 20.6% - 30.6% | 26.5% | 23.9% - 29.1% |  |  |  |
| **Employment status** |  |  |  |  |  |  | **0.4** | **0.3** | **<0.001** |
| Working full-time | 52.1% | 46.1% - 58.6% | 58.7% | 54.2% - 63.7% | 46.0% | 43.4% - 48.7% |  |  |  |
| Working part-time | 13.9% | 8.0% - 20.5% | 12.3% | 7.6% - 17.2% | 14.1% | 11.5% - 16.8% |  |  |  |
| Not working | 34.0% | 28.0% - 40.6% | 29.0% | 24.3% - 33.8% | 39.9% | 37.3% - 42.5% |  |  |  |
| **Poverty level** |  |  |  |  |  |  | **0.076** | **0.3** | **0.2** |
| Below poverty level | 13.9% | 10.2% - 18.1% | 7.4% | 5.3% - 9.9% | 10.2% | 8.7% - 11.5% |  |  |  |
| Above poverty level | 86.1% | 82.3% - 90.3% | 92.6% | 90.6% - 95.2% | 89.8% | 88.5% - 91.3% |  |  |  |
| **COVID-19 vaccination status<sup>†</sup>** |  |  |  |  |  |  | **<0.001** | **<0.001** | **<0.001** |
| Vaccinated | 61.0% | 55.2% - 67.3% | 77.2% | 73.5% - 81.4% | 89.4% | 87.9% - 90.8% |  |  |  |
| Not vaccinated | 39.0% | 33.2% - 45.3% | 22.8% | 19.0% - 26.9% | 10.6% | 9.1% - 12.1% |  |  |  |
| **Total no. of Symptoms lasting 4 or more weeks** |  |  |  |  |  |  | **0.04** | **<0.001** | **<0.001** |
| No Symptoms (0) | 39.4% | 33.0% - 45.8% | 52.2% | 47.3% - 57.1% | 77.7% | 75.7% - 79.6% |  |  |  |
| Any Symptoms (1+) | 60.6% | 55.1% - 67.0% | 47.8% | 43.1% - 52.8% | 22.3% | 20.3% - 24.4% |  |  |  |
| One Symptom (1) | 23.5% | 17.2% - 30.0% | 16.9% | 12.1% - 21.9% | 10.0% | 8.2% - 12.0% |  |  |  |
| Two Symptom (2) | 8.3% | 1.9% - 14.7% | 5.6% | 0.8% - 10.5% | 4.3% | 2.4% - 6.3% |  |  |  |
| Three or More Symptoms (3+) | 28.8% | 22.5% - 35.3% | 25.3% | 20.4% - 30.2% | 8.0% | 6.1% - 10.0% |  |  |  |

\* p-value for weighted Wald chi-square test; all p-values <0.05 indicate significant differences.
° Statistical weighting was used to align the sample with U.S. population distributions, adjusting for gender, age, and education. Weights were designed to match the U.S. Census' American Community Survey (ACS) proportions for these variables.
† For those who reported testing positive for SARS-CoV-2, vaccinated respondents received two doses of a two-dose primary series or one dose of a single dose primary series prior to the date they reported first testing positive. Negative respondents reported whether they were vaccinated (received two doses of a two-dose series or one dose of a single dose series) at the time they completed the survey. Respondents who did not complete their primary series (received one dose of a two-dose series) were excluded from these values (n = 42).

Approximately two-thirds of respondents in each group were White (68.2% among those testing positive during the Delta period, 63.2% among those positive during early-Omicron period, and 62.6% among those who always tested negative). A little more than half of respondents in each group were female (54.5% among those testing positive during Delta predominant period, 56.3% among those testing positive during early-Omicron predominant period, and 50.5% among those who always tested negative). Respondents who tested positive during Delta predominant period were more likely to live in the Midwest than the other test groups (29.5% compared to 17.2% in those testing positive during early-Omicron predominant period and 18.1% among the tested negative group, p<0.05). Respondents who reported testing positive during the Delta predominant period were younger and less likely to be vaccinated prior to infection than respondents who reported testing positive during the Omicron predominant period and reported testing negative (*p*<0.05). Respondents testing positive during Delta predominant period had lower educational attainment and were less likely to be vaccinated prior to infection than respondents testing positive during early-Omicron predominant period (*p*<0.05). Those testing positive during early-Omicron predominant period were younger, had higher educational attainment, were more likely to report working full-time, and were less likely to be vaccinated than negative respondents (*p*<0.05).

### Characteristics of those experiencing ongoing symptoms

Symptoms lasting four or more weeks were reported by 60.6% of respondents who tested positive during the Delta predominant period, 47.8% of those testing positive during the early-Omicron predominant period, and 22.3% of those who reported testing negative (Table 1). More than 25% of respondents testing positive experienced three or more ongoing symptoms which was greater than the rate among the negative respondents (28.8% and 25.3% among those positive during the Delta- and early-Omicron predominant periods, respectively, compared to 8.0% among the negative respondents). Among respondents testing positive and negative, fatigue was the most commonly reported symptom (32.6% among respondents testing positive during Delta predominant period, 26.3% among those testing positive during early-Omicron predominant period, 8.2% among negative respondents; p <0.05) (Table 2). Respondents testing positive during Delta predominant period reported ongoing changes in smell/taste and shortness of breath more often than those testing positive during early-Omicron predominant period (39.7% vs. 14.3% and 17.7% vs. 10.0%, respectively; *p*<0.05).

**Table 2.** Prevalence of ongoing symptoms lasting 4 weeks or longer among respondents by testing status, weighted°.

|  | First testing<br>positive during<br>Delta | First testing<br>positive<br>during<br>Omicron | Reporting<br>only testing<br>negative | Absolute<br>difference<br>test positive<br>(Delta) – test<br>positive<br>(Omicron) | 95% CI | Absolute<br>difference<br>test positive<br>(Delta) – test<br>negative | 95% CI | Absolute<br>difference test<br>positive<br>(Omicron) –<br>test negative | 95% CI |
| --- | --- | --- | --- | --- | --- | --- | --- | --- | --- |
|  | **n = 257** | **n = 460** | **n = 1,705** |  |  |  |  |  |  |
| **Gastrointestinal (GI)**<br>**Symptom** |  |  |  |  |  |  |  |  |  |
| Yes | 13.1% † | 7.8% | 4.2% ‡ | 5.3% | 0.5% - 10.1% | 8.9% | 4.7% - 13.1% | 3.6% | 1.0% - 6.2% |
| Diarrhea | 8.8% † | 3.0% | 1.8% ‡ | 5.8% | 2.0% - 9.6% | 7.0% | 3.5% - 10.5% | 1.2% | -0.5% - 2.9% |
| Nausea/vomiting | 8.2% † | 3.1% § | 1.3% ‡ | 5.1% | 1.4% - 8.8% | 6.9% | 3.5% - 10.3% | 1.8% | 0.1% - 3.5% |
| Stomach pain | 4.2% | 4.2% | 2.2% | 0.0% | -3.1% - 3.1% | 2.0% | -0.6% - 4.6% | 2.0% | 0% - 4.0% |
| **Neurological**<br>**Symptom** |  |  |  |  |  |  |  |  |  |
| Yes | 40.3% † | 31.7% § | 14.0% ‡ | 8.6% | 1.3% - 16.0% | 26.3% | 20.1% - 32.5% | 17.7% | 13.1% - 22.3% |
| Difficulty<br>thinking/brain fog | 12.1% | 12.1% § | 3.4% ‡ | 0.0% | -5.0% - 5.0% | 8.7% | 4.6% - 12.8% | 8.7% | 5.6% - 11.8% |
| Fatigue | 32.6% † | 26.3% § | 8.2% ‡ | 6.3% | -0.7% - 13.3% | 24.4% | 18.5% - 30.3% | 18.1% | 13.9% - 22.3% |
| Headache | 16.7% | 14.2% § | 4.1% ‡ | 2.5% | -3.1% - 8.1% | 12.6% | 7.9% - 17.3% | 10.1% | 6.8% - 13.4% |
| Post-exertional<br>malaise | 5.6% | 6.2% § | 0.7% ‡ | 0.6% | -4.2% - 3.0% | 4.9% | 2.1% - 7.7% | 5.5% | 3.3% - 7.7% |
| Problems sleeping | 9.4% | 6.9% | 5.8% | 2.5% | -1.8% - 6.8% | 3.6% | -0.1% - 7.3% | 1.1% | -1.5% - 3.7% |
| **Respiratory/Cardiac**<br>**Symptom** |  |  |  |  |  |  |  |  |  |
| Yes | 27.7% | 31.2% § | 8.8% ‡ | 3.5% | -10.4% - 3.4% | 18.8% | 13.3% - 24.5% | 22.4% | 18.0% - 26.8% |
| Chest pain | 6.6% | 6.1% § | 1.3% ‡ | 0.5% | -3.7% - 4.7% | 5.3% | 2.2% - 8.4% | 4.8% | 2.6% - 7.1% |
| Cough | 17.6% | 22.9% § | 5.0% ‡ | 5.3% | -12.2% - 1.6% | 12.6% | 7.8% - 17.4% | 17.9% | 13.9% - 21.9% |
| Heart palpitations | 5.7% | 3.4% | 1.6% ‡ | 2.3% | -1.3% - 5.9% | 4.1% | 1.2% - 7.0% | 1.8% | 0% - 3.6% |
| Shortness of breath | 17.7% † | 10.0% § | 3.2% ‡ | 7.7% | 1.8% - 13.6% | 14.5% | 9.8% - 19.2% | 6.8% | 3.9% - 9.7% |
| Sore throat | 9.9% | 11.4% § | 1.9% ‡ | 1.5% | -6.8% - 3.8% | 8.0% | 4.3% - 11.7% | 9.5% | 6.5% - 12.5% |
| **Other Symptom** |  |  |  |  |  |  |  |  |  |
| Yes | 42.8% † | 27.5% § | 11.4% ‡ | 15.3% | 8.0% - 22.6% | 31.4% | 25.2% - 37.6% | 16.1% | 11.8% - 20.5% |
| Change in mood | 3.9% | 5.0% | 1.8% | 1.1% | - 4.2% - 2.0% | 1.8% | -0.4% - 4.6% | 3.2% | 1.11% - 5.3% |
| Change in smell/taste | 29.7% † | 14.3% § | 1.0% ‡ | 15.4% | 9.0% - 21.8% | 28.7% | 23.1% - 34.3% | 13.3% | 10.1% - 16.5% |
| Fever | 15.3% | 10.9% § | 1.5% ‡ | 4.4% | -8.4% - 9.6% | 13.8% | 9.4% - 18.2% | 9.4% | 6.5% - 12.3% |
| Hair loss | 9.1% † | 6.6% § | 1.5% ‡ | 2.5% | -1.7% - 6.7% | 7.6% | 4.0% - 11.2% | 5.1% | 2.8% - 7.4% |
| Joint/muscle pain | 14.7% | 10.9% | 8.2% ‡ | 3.8% | -1.4% - 9.0% | 6.5% | 2.0% - 11.0% | 2.7% | -0.4% - 5.8% |

° Statistical weighting was used to align the sample with U.S. population distributions, adjusting for gender, age, and education. Weights were designed to match the U.S. Census' American Community Survey (ACS) proportions for these variables.
† p-value for pairwise comparison of proportions test between positives during Delta predominant period and positives during Omicron predominant period <0.05, differences between those first testing positive during Delta predominant period and those first testing positive for SARS-CoV-2 during Omicron predominant period.
‡ p-value for pairwise comparison of proportions test between positives during Delta predominant period and Negatives <0.05, differences between those first testing positive during Delta predominant period and those receiving a negative SARS-CoV-2 test result.
§ p-value for pairwise comparison of proportions test between positives during Omicron predominant period and Negatives <0.05, differences between those first testing positive during Omicron predominant period and those receiving a negative SARS-CoV-2 test result.

### Healthcare utilization

Report of healthcare utilization was higher among the positive groups than those who reported testing negative (Table 3). Among respondents positive during Delta predominant period, 57.7% reported utilizing some health services for their ongoing symptoms, compared to the 35.9% of negative respondents (p<0.05). Respondents positive during the Delta predominant period were more likely to be hospitalized and treated in an intensive care unit (ICU) than those positive during the early-Omicron predominant period (9.6% hospitalized compared to 0.4% and 2.5% treated in ICU compared to 0%, respectively; p<0.05).

**Table 3.** Healthcare utilization for ongoing symptoms lasting 4 weeks or longer by testing status, weighted°.

|  | First testing<br>positive during<br>Delta | 95% CI | First<br>testing<br>positive<br>during<br>Omicron | 95% CI | Testing<br>negative | 95% CI | Delta v.<br>Omicron | Delta v.<br>Negative | Omicron<br>v.<br>Negative |
| --- | --- | --- | --- | --- | --- | --- | --- | --- | --- |
|  | **n = 156** |  | **n = 185** |  | **n = 317** |  |  |  |  |
| **Utilized healthcare for ongoing symptoms** |  |  |  |  |  |  |  |  |  |
| Yes | 57.7% | 50.3% - 65.9% | 45.5% | 38.7% - 52.6% | 35.6% | 30.7% - 40.7% | **0.06** | **<0.001** | **0.05** |
| **Received outpatient care<sup>x</sup>** |  |  |  |  |  |  |  |  |  |
| Yes | 53.1% | 45.7% - 61.3% | 45.5% | 38.7% - 52.6% | 34.5% | 29.8% - 39.7% | **0.2** | **0.001** | **0.03** |
| Saw health professional | 33.2% | 26.5% - 41.1% | 32.8% | 26.5% - 39.4% | 29.6% | 24.9% - 34.3% | >0.9 | 0.5 | 0.5 |
| Went to urgent/emergency care | 24.8% | 18.6% - 31.6% | 14.8% | 10.5% - 19.8% | 8.6% | 6.1% - 11.5% | 0.05 | <0.001 | 0.06 |
| **Received inpatient care<sup>y</sup>** |  |  |  |  |  |  |  |  |  |
| Yes | 10.0% | 6.2% - 14.7% | 0.4% | 0% - 1.4% | 2.5% | 1.2% - 4.0% | **0.008** | **0.04** | **0.03** |
| Hospitalized | 9.6% | 5.8% - 14.2% | 0.4% | 0% - 1.4% | 1.3% | 0.5% - 2.5% | 0.01 | 0.02 | 0.2 |
| Treated in ICU | 2.5% | 0.6% - 4.8% | 0.0% | 0% - 0.8% | 1.4% | 0.5% - 2.6% | 0.04 | 0.4 | 0.05 |
| Given breathing tube/ventilator | 1.5% | 0.3% - 3.3% | 0.0% | 0% - 0.8% | 0.8% | 0.2% - 1.8% | 0.2 | 0.5 | 0.12 |

<sup>°</sup> Statistical weighting was used to align the sample with U.S. population distributions, adjusting for gender, age, and education. Weights were designed to match the U.S. Census' American Community Survey (ACS) proportions for these variables.
\* p-value for weighted Wald chi-square test; all p-values <0.05 indicate significant differences.
<sup>x</sup> Outpatient care included seeing a doctor, nurse, or another health professional or receiving emergency or urgent care. Respondents could have reported utilizing more than one type of outpatient care service
<sup>y</sup> Inpatient care included being hospitalized, being treated in an intensive care unit (ICU) or being placed on a ventilator or given a breathing tube. Respondents could have reported utilizing more than one type of inpatient care service

### Factors associated with ongoing symptoms lasting ≥ 4 weeks

Respondents positive during the Delta- and early-Omicron predominant periods had greater odds of reporting an ongoing symptom lasting four or more weeks relative to respondents who tested negative (adjusted odds ratio [aOR], 5.94; 95% CI, 4.48-7.90; aOR, 3.13; 95% CI, 2.51-3.91, respectively) (Table 4). Socio-demographic factors associated with increased odds of reporting ongoing symptoms lasting four or more weeks among all respondents were being female, and report of being unemployed. When examining positive respondents only, the odds of reporting an ongoing symptom were lower among those testing positive during the early-Omicron predominant period than those testing positive during the Delta predominant period and were higher among those who were female (aOR, 0.53; 95% CI, 0.38 – 0.72; aOR, 1.36; 95% CI, 1.00-1.85, respectively).

**Table 4.** Logistic regression analyses of risk factors associated with ongoing symptoms lasting four weeks or longer.

|  | Model 1: Respondent reporting ongoing symptoms (n = 2,421) |  |  |  | Model 2: Respondents testing positive and reporting ongoing symptoms (n = 716) |  |  |  |
| --- | --- | --- | --- | --- | --- | --- | --- | --- |
|  | Unadjusted OR | 95% CI | Adjusted OR | 95% CI | Unadjusted OR | 95% CI | Adjusted OR | 95% CI |
| **SARS-CoV-2 test status** |  |  |  |  |  |  |  |  |
| Tested negative | 1.00 | Reference | 1.00 | Reference | N/A | N/A | N/A | N/A |
| Positive during Delta | 5.53 | 4.20 - 7.31 | 5.94 | 4.48 - 7.90 | 1.0 | Reference | 1.0 | Reference |
| Positive during Omicron | 2.93 | 2.36 - 2.64 | 3.13 | 2.51 - 3.91 | 0.53 | 0.39 - 0.72 | 0.53 | 0.38 - 0.72 |
| **Age Group (years)** |  |  |  |  |  |  |  |  |
| 18-39 | 1.00 | Reference | 1.00 | Reference | 1.00 | Reference | 1.00 | Reference |
| 40-59 | 1.00 | 0.79 - 1.27 | 1.01 | 0.79 - 1.31 | 1.24 | 0.86 - 1.78 | 1.25 | 0.86 - 1.82 |
| >= 60 | 1.01 | 0.80 - 1.27 | 1.08 | 0.82 - 1.42 | 1.22 | 0.82 - 1.82 | 1.13 | 0.73 - 1.76 |
| **Sex** |  |  |  |  |  |  |  |  |
| Male | 1.00 | Reference | 1.00 | Reference | 1.00 | Reference | 1.00 | Reference |
| Female | 1.34 | 1.13 - 1.59 | 1.28 | 1.06 - 1.55 | 1.34 | 1.00 - 1.80 | 1.36 | 1.00 - 1.85 |
| **Employment status** |  |  |  |  |  |  |  |  |
| Working full time | 1.00 | Reference | 1.00 |  | 1.00 | Reference | 1.00 | Reference |
| Working part-time | 1.00 | 0.75 - 1.32 | 1.03 | 0.75 - 1.40 | 0.95 | 0.59 - 1.52 | 0.89 | 0.54 - 1.46 |
| Not working | 1.28 | 1.06 - 1.53 | 1.48 | 1.17 - 1.87 | 1.36 | 0.98 - 1.89 | 1.29 | 0.88 - 1.89 |

° Statistical weighting was used to align the sample with U.S. population distributions, adjusting for gender, age, and education. Weights were designed to match the U.S. Census' American Community Survey (ACS) proportions for these variables.

## Discussion

In this representative sample of non-institutionalized adults in the United States, prior SARS-CoV-2 infection was associated with a higher prevalence of ongoing symptoms lasting 4 or more weeks than those who always tested negative. The prevalence of ongoing symptoms was significantly lower among those who tested positive during early-Omicron than those who tested positive during the Delta period (47.8% compared to 60.6%, respectively). Those who tested positive during the Delta period reported gastrointestinal and neurological symptoms than the early-Omicron group.

Studies have similarly shown ongoing symptoms are more prevalent among individuals previously infected during the Delta and Omicron periods than negative controls^19,20^. Our findings demonstrated the odds of reporting an ongoing symptom were lower following a positive test during the Omicron period than the Delta period. A case-control observational study similarly found those testing positive during Omicron had reduced odds of long COVID compared to those testing positive during Delta^13^. Doll et al.’s study including adults who were unvaccinated prior to infection also observed a significant reduction in the occurrence of ongoing symptom among Omicron cases compared to Delta cases^21^.

The most common ongoing symptoms reported in our study were fatigue, cough, and change in smell/taste. A cross-sectional internet survey, whose study design was similar to the current study, found fatigue and loss of smell were the most common ongoing symptoms reported by those infected during the Delta and Omicron periods^5^. The Norwegian study mentioned previously similarly observed a 20-30% increase in the estimated risk of ongoing fatigue among individuals testing positive in either the Delta or Omicron periods compared to the negative individuals^19^.

Respondents who tested positive during the early-Omicron period were less likely to report change in smell/taste and nausea/vomiting than those who tested positive during the Delta period. Results from the INSPIRE study similarly demonstrate lower odds of loss of taste/smell and nausea/vomiting in those in the Omicron cohort when compared to the Delta cohort^12^. Females were more likely to report an ongoing symptom than males in our study. Similarly, Perlis et al.’s cross-sectional internet survey found a significant association between female gender and ongoing symptoms^5^.

Those who tested positive during the Delta period were more likely to be hospitalized and treated in the intensive care unit for their ongoing symptom(s) than those who tested positive during the early-Omicron period. These differences in healthcare utilization might be explained by the clinical severity of the acute COVID-19 case. Studies have compared the clinical severity of COVID-19 based on SARS-CoV-2 variant and have established infection with the Delta variant is more severe than infection with an Omicron variant^9,10,21,22^. This is further supported by data from electronic health records and self-reported surveys further demonstrated more severe cases of acute COVID-19 were associated with post-COVID conditions ^23^.

Although we did not examine the effect in this study, a growing body of research suggest post-COVID conditions are less likely to occur among persons vaccinated prior to SARS-CoV-2 infection. An Israeli study found receiving at least two doses of a COVID-19 vaccine was associated with decreased reports of post-COVID condition^6^. Among Omicron cases in Nehme et al.’s Swiss study, ongoing symptoms were more common among unvaccinated than vaccinated respondents^20^. As individuals continue to vaccinate against new variants, it is important to assess the possible protective effects of vaccination on post-COVID conditions.

### Limitations and Strengths

This cross-sectional study has a number of limitations. First, respondents who participated in this survey may not be representative of all non-institutionalized U.S. adults. Further, those who did not report ever testing for COVID-19 and who were excluded from the analysis may be different from those who did test. Second, self-report of COVID-19 testing, vaccination, and ongoing symptoms may be subject to response bias and potential misclassification of SARS-CoV-2 test result. If respondents who reported testing negative had a false negative test result, this attenuated the results towards the null. The survey did not capture timing of vaccination so it is possible that vaccination status for some respondents may not have reflected vaccination status at the time of their infection due to lack of temporal clarity. Finally, the length of time between reporting testing positive for COVID-19 and completion of the survey was 6 months or longer for those who tested positive during the Delta period. Since the survey asks about ongoing symptoms experienced in the past week, the duration of symptoms among those positive during the Delta and early-Omicron periods may differ. Including measures of ongoing symptom severity and the severity of the initial SARS-CoV-2 infection may have also provided additional information about risk factors for post-COVID conditions.

Despite these limitations, this study had several strengths. Our study had a high response rate. Questions included in the survey were standardized, including the list of ongoing symptoms. We included those who reported at least one positive test and those who always tested negative for SARS-CoV-2 in our sample. Including respondents who tested negative and positive in the analysis allowed us to estimate the prevalence of these ongoing symptoms among these populations, enabling us to estimate the percentage of symptoms that might be attributable to SARS-CoV-2 among adults.

While report of ongoing symptoms lasting 4 weeks or longer that are associated with post-COVID conditions may have declined during the Omicron variant period, a large percentage of adults who tested positive were still impacted. As new Omicron variants emerge it is important to continue evaluating the burden of post-COVID conditions. Continuing to assess the impact of post-COVID conditions will provide guidance to clinicians, public health professionals and health care organizations in addressing ongoing health care needs.

## Data Availability

The data underlying this study were provided by Porter Novelli Public Services under the terms of a research agreement. Inquiries regarding access to the data should be made directly to Porter Novelli.

## Acknowledgements

DE, PL, and SS contributed to the conceptualization of this study. DE and EA analyzed the data, designed, and drafted the figures and tables. All authors interpreted the results and DE prepared the initial manuscript draft. PL, LF, TF, EA, and SS provided necessary revisions prior to submission for publication. All authors contributed to, reviewed, and approved the final draft of this paper.

There are no conflicts of interest between the authors listed in this manuscript. This study was not funded using financial support from grant awards.

## Appendix

|  |  |
| --- | --- |
| Change in mood | Other |
| Change in smell or taste | Other |
| Chest pain/pressure | Respiratory/Cardiac symptoms |
| Cough | Respiratory/Cardiac symptoms |
| Diarrhea | GI symptoms |
| Difficulty thinking clearly, concentrating, forgetfulness, memory loss, or 'brain fog' | Neurological symptoms |
| Fatigue/tired/weakness | Neurological symptoms |
| Fever or chills | Other |
| Hair loss | Other |
| Headache | Neurological symptoms |
| Joint/muscle pain | Other |
| Nausea/vomiting | GI symptoms |
| Palpitations (heart racing or pounding) | Respiratory/Cardiac symptoms |
| Post-exertional malaise (worsening of symptoms after even minor physical, mental, or emotional exertion) | Neurological symptoms |
| Problems sleeping | Neurological symptoms |
| Shortness of breath/breathlessness | Respiratory/Cardiac symptoms |
| Sore throat | Respiratory/Cardiac symptoms |
| Stomach pain | GI symptoms |

